# Post natal Care services utilization and satisfaction of Women in Afar Region, Awash Fentale District, Ethiopia

**DOI:** 10.1101/2024.01.31.24302088

**Authors:** Getachew Weldeyohannes, Thanyani Gladys

## Abstract

**Back ground:** The percentage of women receiving a postnatal checkup within two days of delivery is higher in urban areas than in rural areas. In Afar region it was 11.6%, which is the lowest among all regional states in Ethiopia

**Aim:** The aim of this study is to assess the post natal care services utilizations and satisfactions of Women in Afar Region, Awash Fentale District.

**METHOD:** A quantitative, descriptive, cross-sectional study design was used for this study. Data were collected using a structured questionnaire administered to 422 women aged 15 to 49 years through a stratified sampling technique. Data was entered, analyzed and interpreted using SPSS computer program. Binary logistic regression model was used to identify the factors that influence maternal health care services utilization.

**Result:** This study revealed that 43% of the respondents were not satisfied with the care and attention given by the health care providers and approximately 52% of the respondents were not satisfied with the cleanliness of the health facilities. Regardless of the level or degree of income, husbands’ income was statistically significant and earning less income among husbands might lead women to have less PNC services utilization. Another independent variable that showed statistical significance was watching television .Women who never watched television were 58% less likely to use PNC services compared to women who watched television every day (AOR=0.414and p-value=0.008), which is less than p=0.05.

**Conclusion:** husbands’ income and watching television were statistically significant and earning less income among husbands might lead women to have less PNC services utilization. Much emphasis should also be given to care, attention of mothers and the cleanliness of the healthcare facilities in Awash Fentale woreda.

## Introduction

The percentage of women receiving a postnatal checkup within two days of delivery is higher in urban areas than in rural areas. The percentage of women who had a postnatal checkup in the first two days after birth in Afar Region was 11.6%, which is the lowest among all regional states in Ethiopia (1). A similar study conducted in Swaziland indicated that the utilization of PNC services was high among women who have had less than six children. The rate of PNC services utilization was low among women who have had six or more children (16.8%) (2). According to Atiya (3), the main factors identified as influencing satisfaction and dissatisfaction were caregivers and client interaction, the characteristics of the setting, the involvement of clients in the caring process, the nurses’ perception of client characteristics, the outcome of labor for both mother and baby, the acceptability of alternative places for delivery, and the respondents’ expectations and perceptions of hospital delivery. Of all these factors, however, caregiver attitude was seen as the strongest factor in determining maternal satisfaction with care”.

## Material and Method

### Study area, period and design

The study was conducted in Awash Fentale district, in the Afar Region of North East of Ethiopia, from January to March 2020. Awash Fentale is found in Zone 3 of the Afar National Regional State, Ethiopia, which is 200km from Addis Ababa.

The total population of Awash Fentale distric for the year 2017 was 37,219 populations, of which 17,880 are women and 19,339 are men. For this study the researcher used a quantitative, descriptive, cross-sectional design and a stratified random sampling was applied. In the stratified sampling, the sampling frame is divided into homogeneous and non-overlapping subgroups (called “strata”), and a simple random sample is drawn within each subgroup. The variables commonly used for stratification was ethnicity. This Stratification ensured that all levels of the identified variables were adequately represented in the sample. Study subjects were selected by simple random sampling from each stratum.

### Sample size determination and procedure

The sample size required for this study was determined using a single population proportion formula with the following assumptions; the proportion of postnatal care utilization in Awash Fentale district was assumed to be 50% (p = 0.5) level of significance 5% (α = 0.05), Z α/2 = 1.96, absolute precision or margin of error 5% (d = 0.05) and non-response rate 10%. Using the formula n = [Z α/2*p(1-p)]/d^2^ = 1.96^2^*0.5 (1–0.5)/0.05^2^, the sample size n was calculated to be 380. Finally, considering a 10% contingency rate for non-responses, the final sample size was calculated as 422.

### Data collection Procedure

Data were collected using standardized structured questionnaires. The data were collected using questionnaires developed and included all the relevant variables to meet the objectives of the study. Prior to the development of the questionnaires, similar studies focusing on factors that influence the utilization of maternal healthcare services were reviewed and a few modifications were made to the prepared questionnaires to address the set objectives. Before the actual data collection, the data collectors were provided with the location and the number of women to interview based on the proportion of their ethnicity. The households of the respondents, regardless of their place of delivery, were obtained from the Maternal and Child Health (MCH) Department of Awash Health Centre. All women aged 15 to 49 years who gave birth within the past two years and who had been living there for at least one year (but not visitors) were eligible for the interview. The data collectors were required to do only one revisit and those households missed during the second visit were assumed as non-respondents. The study respondents were randomly selected from their ethnic strata and the number of respondents in each ethnic group who were interviewed by the data collectors were proportionally allocated by the researcher. During the data collection, the researcher consulted the heads of Awash Fentale Health Office and Awash Fentale Health Centre and some community members to make the data-collection process more effective. In this study, verbal face-to-face interviews were administered using structured questionnaires. The questionnaires were developed in English and translated into Amharic and Afar language and further explanations were given for better understanding of enumerators and respondents.

### Data quality control

The questionnaires were prepared separately according to the nature of participants. Data collectors and supervisors were recruited and received training for three consecutive days. The questioning tool was pretested prior to the actual data collection to check its appropriateness and the familiarity of the data collectors with the tool. The data collectors collected the data under continuous supervision and guidance by the supervisors and investigators.

### Ethics

Ethical clearance certificate was obtained from the University of South Africa (Ref:HSHDC/594/2017) and from the Afar National Regional State Bureau of Health (Ref:A/HIC/719/10). A permission letter to contact the list of women was obtained from Awash Fentale Health Centre which has the authoritative power and responsibility of leading the healthcare facilities.

A written informed consent form was designed and was signed by each respondent before completing the questionnaire: The researcher respected the rights of the respondents to abstain from participation or to withdraw consent to participate at any time without reprisal. The participants were informed that the information they provided might not be of direct benefit to them but was extremely important to inform policy makers and program designers for stimulating discussion about the formulation of appropriate measures to address the factors that influence maternal healthcare services utilization by women in Awash Fentale woreda. Case identification numbers instead of actual names were used to ensure anonymity. Data was locked in the box and kept in the office of the manager of the health center for safety from disclosure to unauthorized persons. Stratified and simple random sampling procedure was used to ensure justice and avoid discrimination of participants. Data collectors were well orientated and obtained data from respondents; homes to avoid social and economic harm.

### Validity and reliability

To ensure external validity, simple random sampling of respondents was done and the sample size was large enough to draw conclusions to generalize findings to other populations. The items on the questionnaire were tested to ensure reliability.

### Data analysis

The researcher reviewed the data for logical consistencies and completeness before making data entries. Data were entered into SPSS and cleaned also using SPSS. The description of the study population was done by analyzing the distribution of the respondents by the variables in terms of frequencies and percentages. Chi-square tests were conducted to assess any association and to measure the strength of association between independent and dependent variables. Similarly, binary logistic regression was applied to assess any association and the strength of the association between dependent and independent variables.

## Result

### Demographic characteristics of respondents

The overall response rate was 422 (100%). Of the total respondents, 156 (36.9%) were illiterate, 228 (54%) were from Afar ethnic group, 207 (49%) were housewives, 275 (65%) were Muslims, 406 (96.2%) were married; 266 (63%) had an income range between 1501–5000 Birr (which is approximately 51–172USD) per month and 164 (39.8%) were in the age group 25 – 29 years with a mean age of 26.9 years. About 298 (70.6%) had history of 2–5 Pregnancies (Table1). Among the respondents, 56(13.3%) had history of abortion, 76 (18%) still birth and 52(12.3%) had an infant death (Table 1).

**Table 1.** Socio demographic characteristics of respondents in Awash Fentale District, Afar Regional State, Ethiopia.

| Characteristics of respondents | Frequency | Percent |
| --- | --- | --- |
| Age |  |  |
| Mean ± SD | 26.9±5 | |
| 15-19 | 35 | 8.3 |
| 20-24 | 88 | 20.8 |
| 25-29 | 164 | 39.8 |
| 30-34 | 85 | 20 |
| 35-39 | 40 | 9.5 |
| 40-44 | 8 | 1.9 |
| **Educational level** |  |  |
| No education | 156 | 36.9 |
| Primary education | 48 | 11.4 |
| Secondary education | 115 | 27.25 |
| Tertiary education and above | 103 | 24.4 |
| **Ethnicity** |  |  |
| Afar | 228 | 54 |
| Amhara | 93 | 22 |
| Oromo | 42 | 10.4 |
| Welayta | 25 | 5.9 |
| Argoba | 17 | 4 |
| Tigray | 8 | 1.9 |
| Others | 2 | 0.5 |
| **Occupation** |  |  |
| Not employed | 207 | 49.1 |
| Government employee | 115 | 27.3 |
| Self employee | 55 | 13 |
| Private employee | 19 | 4.5 |
| NGO | 2 | 2.1 |
| Unspecified job | 17 | 4 |
| **Religion** |  |  |
| Muslism | 275 | 65 |
| Orthodox Christian | 92 | 22 |
| Protestant | 44 | 10.4 |
| Catholic | 10 | 2.4 |
| Traditional believers | 1 | 0.2 |
| **Marital Status** |  |  |
| Married | 406 | 96.2 |
| Cohabiting | 2 | 0.5 |
| Never married | 4 | 0.9 |
| Widowed | 5 | 1.2 |
| Divorced | 4 | 0.9 |
| **Income** |  |  |
| ≤1500 | 103 | 24 |
| 1501-5000 | 266 | 63 |
| ≥5000 | 53 | 12.6 |

### PNC services utilization

Of the 422 respondents, 234 (55.45%) had PNC visits, while 185(44.4%) did not have any PNC visits. Only three (0.7%) of the women could not remember whether they had PNC visits or not. Figure 4.13 indicates the proportion of women who had PNC visits at a healthcare facility after delivery (see figure 2).

**(a) Barriers to PNC services in Awash Fentale District**
It was indicated in this study that 135 (42.2%) of the women responded they did not access PNC services because they did not experience any complications after delivery while they were at home. The second reason mentioned for not visiting the healthcare facility after delivery was because their family members did not allow them to visit a healthcare facility. Thirty-three (10.3%) of the respondents indicated their husbands’ influence as a barrier. Long distance from the health care facility, not trusting the health care facilities, and the unavailability of quality services were among the major reasons mentioned by the study participants that prevented them from visiting a health care facility (see Table 2).

**Figure 2:**
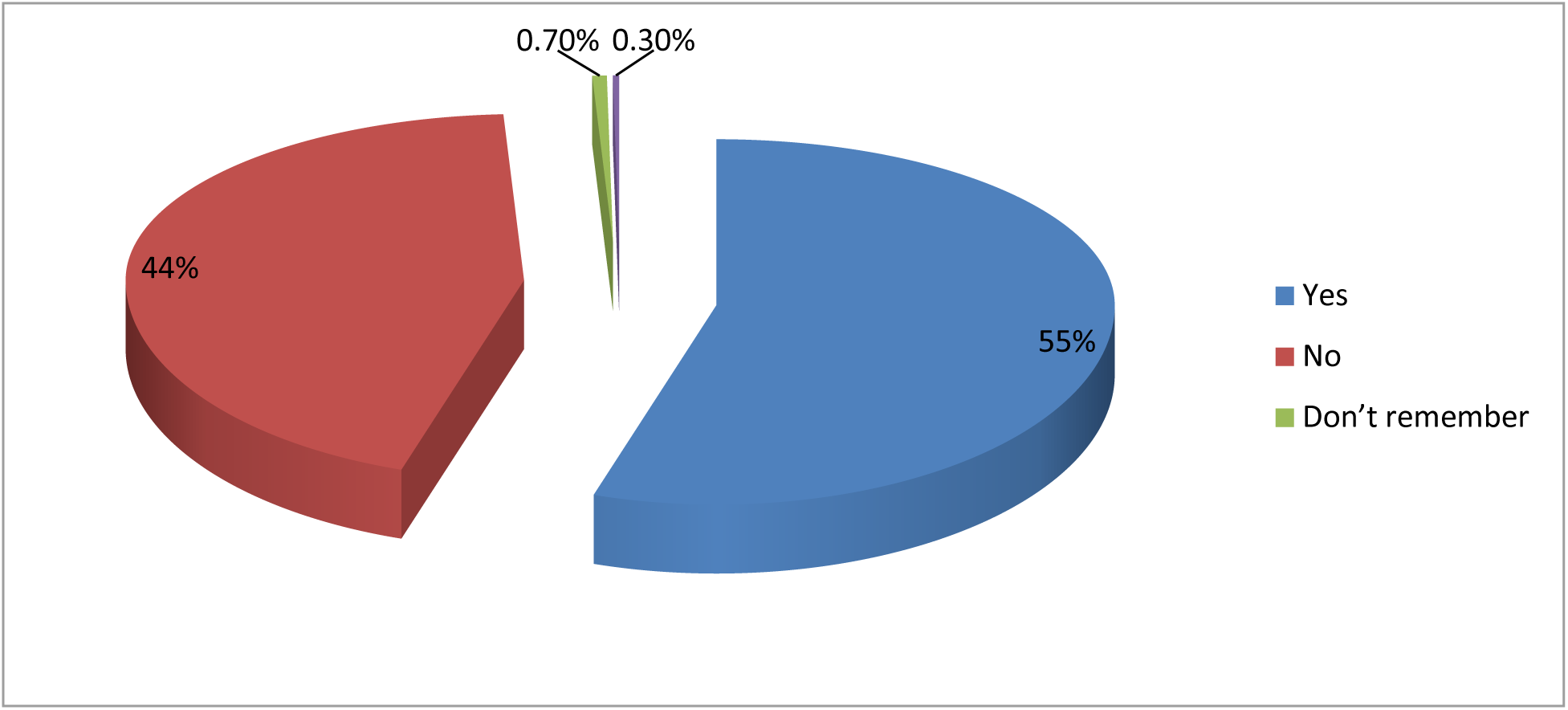
Proportion of women who had PNC services

**Table 2:** Proportion of the responses regarding barriers to accessing PNC services (n=320)

| Reasons not to have PNC | Frequency | Percentage |
| --- | --- | --- |
| High cost of service | 10 | 3.1 |
| Long distance | 22 | 6.9 |
| Long waiting time | 13 | 4.1 |
| No complication | 135 | 42.2 |
| Do not trust healthcare facility | 22 | 6.9 |
| Unavailability of quality service | 22 | 6.9 |
| Husband did not allow it | 33 | 10.3 |
| Family did not allow it | 43 | 13.4 |
| Previous bad experience at a healthcare facility | 5 | 1.56 |
| Little respect from healthcare workers | 15 | 4.7 |
| **Total responses** | **320** | **100.00** |

**Table 3:** Women’s satisfaction with maternal health care services (N=385)

| **Satisfaction** | **Frequency** |  |  | **Percentage** |  |  |
| --- | --- | --- | --- | --- | --- | --- |
|  | **Yes** | **No** | **Do not remember** | **Yes** | **No** | **Do not remember** |
| The care and attention from staff | 200 | 166 | 19 | 52 | 43.1 | 4.9 |
| The attitude of healthcare personnel | 153 | 200 | 32 | 39.7 | 52 | 8.3 |
| The cleanliness of the healthcare facility | 170 | 200 | 15 | 44.1 | 52 | 3.9 |
| The amount of privacy | 135 | 223 | 27 | 35 | 58 | 7 |
| The medication provided | 82 | 248 | 55 | 21.3 | 64.4 | 14.3 |

### Women’s satisfaction with maternal health care services

Respondents who had PNC visits were asked whether they had been satisfied with the care they received. Out of 385 women who responded to this question, 200 (52%) were satisfied with the care and attention they received from the healthcare provider, while 166 (43.1%) responded that they were not satisfied with the care and attention they received from the healthcare provider. Only 32 (8.3%) said that they did not remember. With regard to the attitude of health personnel, 153 (39.7%) of the respondents were satisfied with the attitude of the health personnel in providing maternal health care services, while 200 (52%) were not satisfied. Less than half (n=170; 44%) of the respondents claimed that they were satisfied with the cleanliness of the healthcare facility, while more than half 200 (52%) answered that they were not satisfied with the cleanliness of the healthcare facility. The respondents were also asked about their satisfaction pertaining to the amount of privacy they had and the medication they received. Only 135 (35%) of the respondents were satisfied with the privacy they had and only 82 (21.3%) were satisfied with the medication they received (see Table 2).

### Other health-related issues

In this subsection, aspects including women’s decision making, reading newspapers or magazines, and the frequency of listening to the radio or watching television are presented.

**(a) Decision making** The study participants were asked about their decision-making process while seeking health care. The study found that 158 (37.4) of the women responded that they made decisions regarding seeking health care with their partner, while 141(33.4%) indicated that their husband or partner decided for them, 72 (17%) decided alone regarding their health care and 38 (9%) decided along with another person. For only 13 (3.1%) of the participants, someone else decided for them (see Figure 3).
**(b) Reading newspapers or magazines** Of the 422 respondents, 328 (77.7%) reported that they did not read either newspapers or magazines at all, while 43 (10.2%) read newspapers or magazines less than once a week and 36 (8.5%) read newspapers at least once a week. Only 15 (3.6%) of the participants read either a newspaper or a magazine every day (See Figure 4).
**(c) Frequency of listening to the radio among the study respondents** More than half (n=250; 59.2%) of the participants reported that they never listened to the radio at all. Seventy-two (17%) respondents listened to the radio less than once a week and 53 (12.6%) listened to the radio at least once a week. Only 47(11%) participants listened to the radio almost every day (see figure 5).
**(d) Frequency of watching television** The study revealed that 266 (63%) of the participants watched television every day, while 95 (22.5%) did not watch television at all. Moreover, 35(8.3%) women watched television less than once a week and only 26 (6.2%) women watched television at least once a week (see Figure 6).

**Figure 3:**
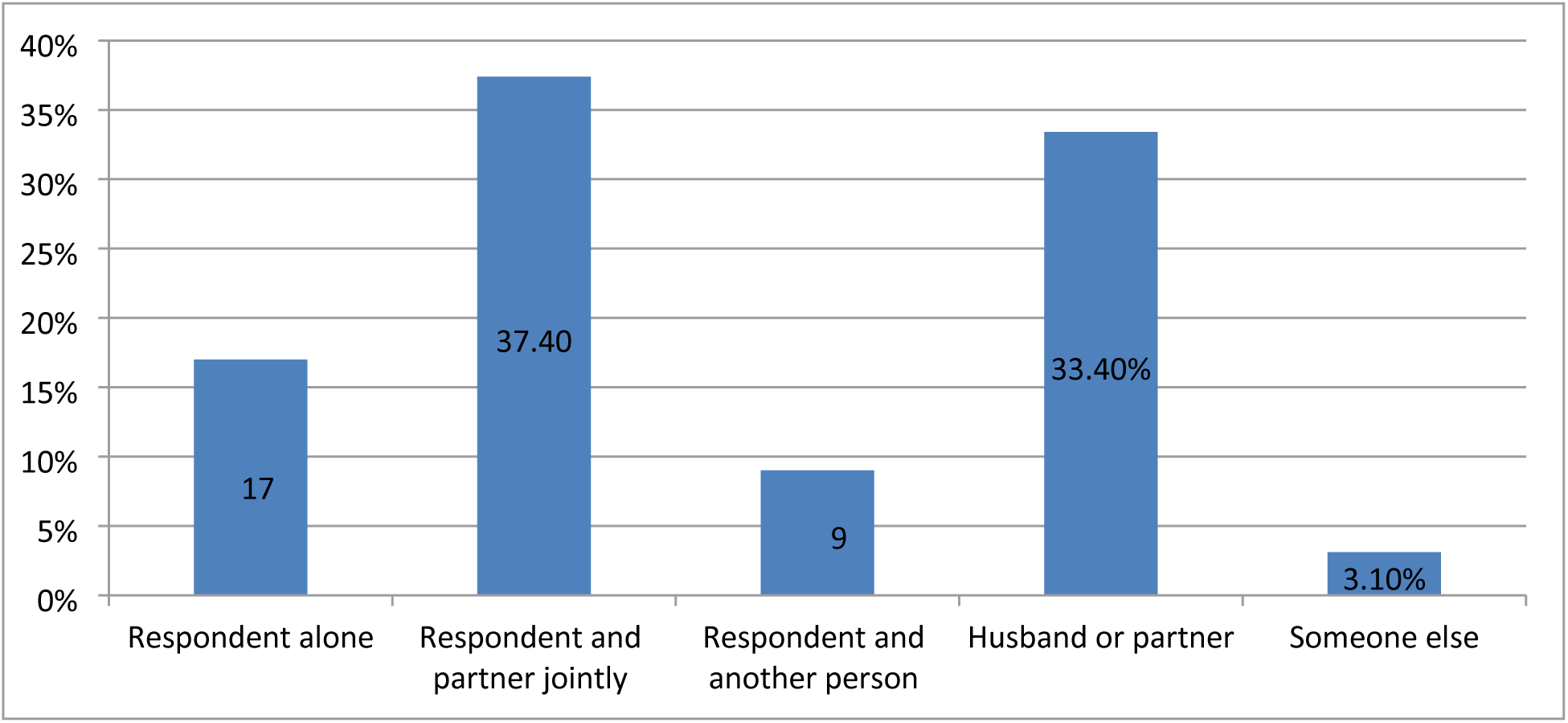
Proportion of respondents regarding their decision making

**Figure 4:**
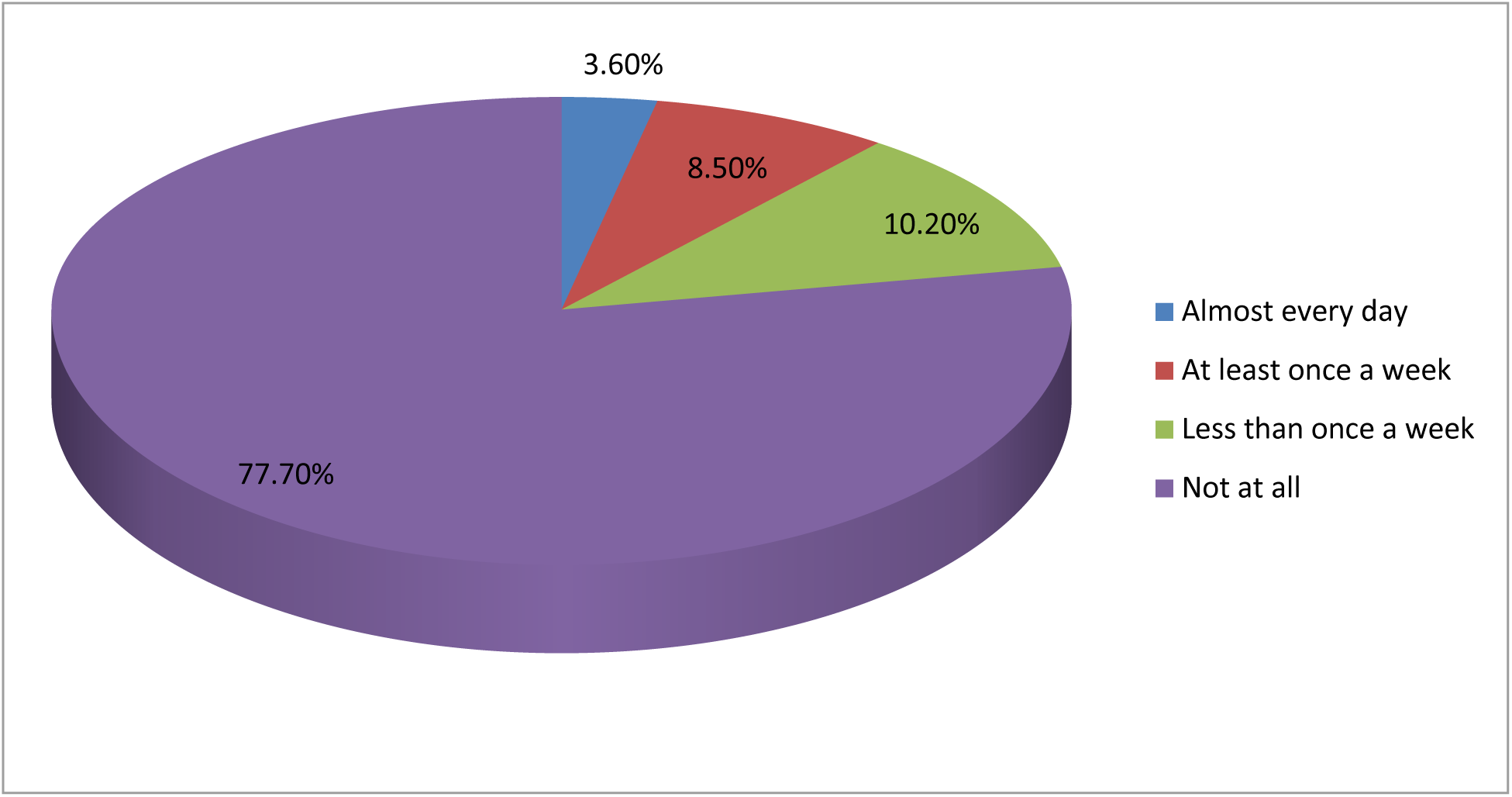
Frequency of reading newspapers or magazines

**Figure 5:**
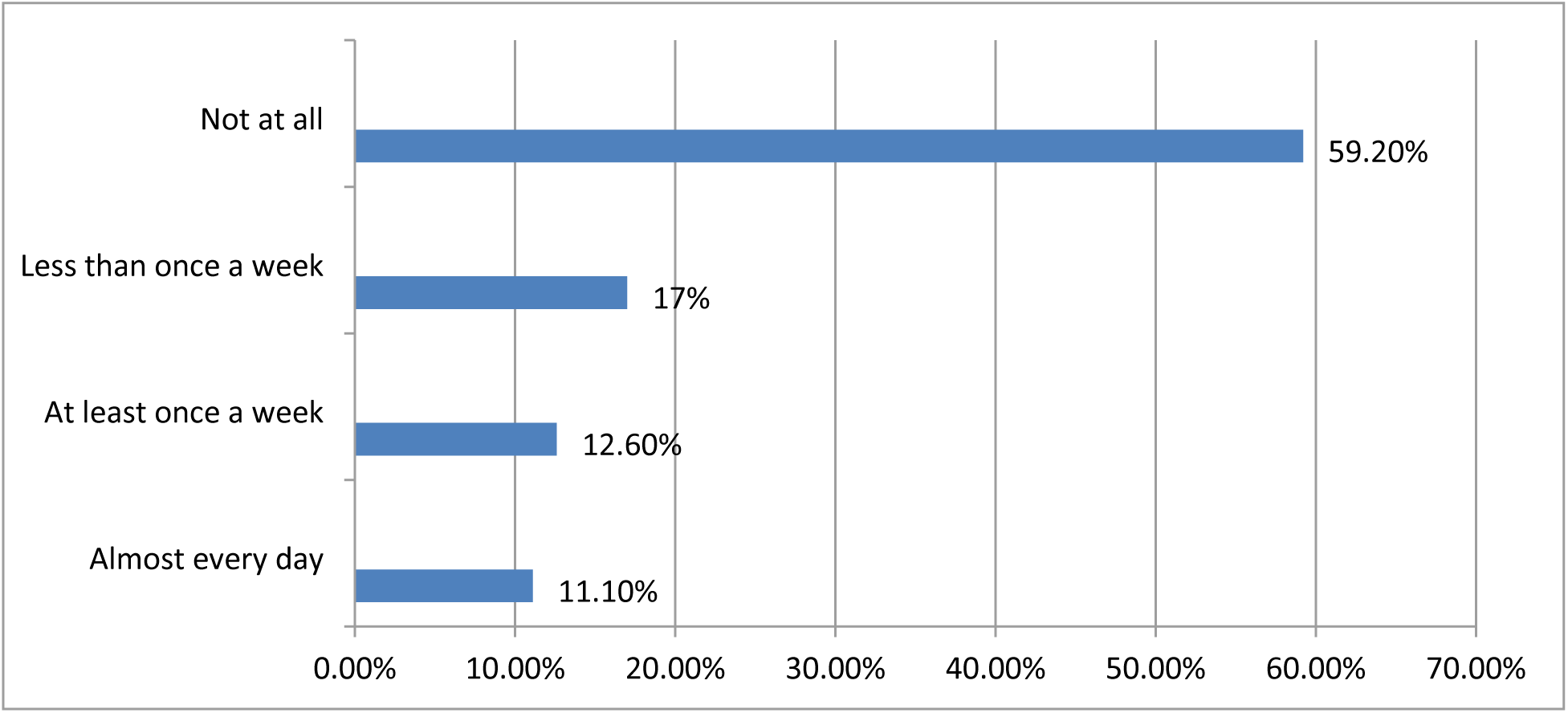
Frequency of listening to the radio

**Figure 6:**
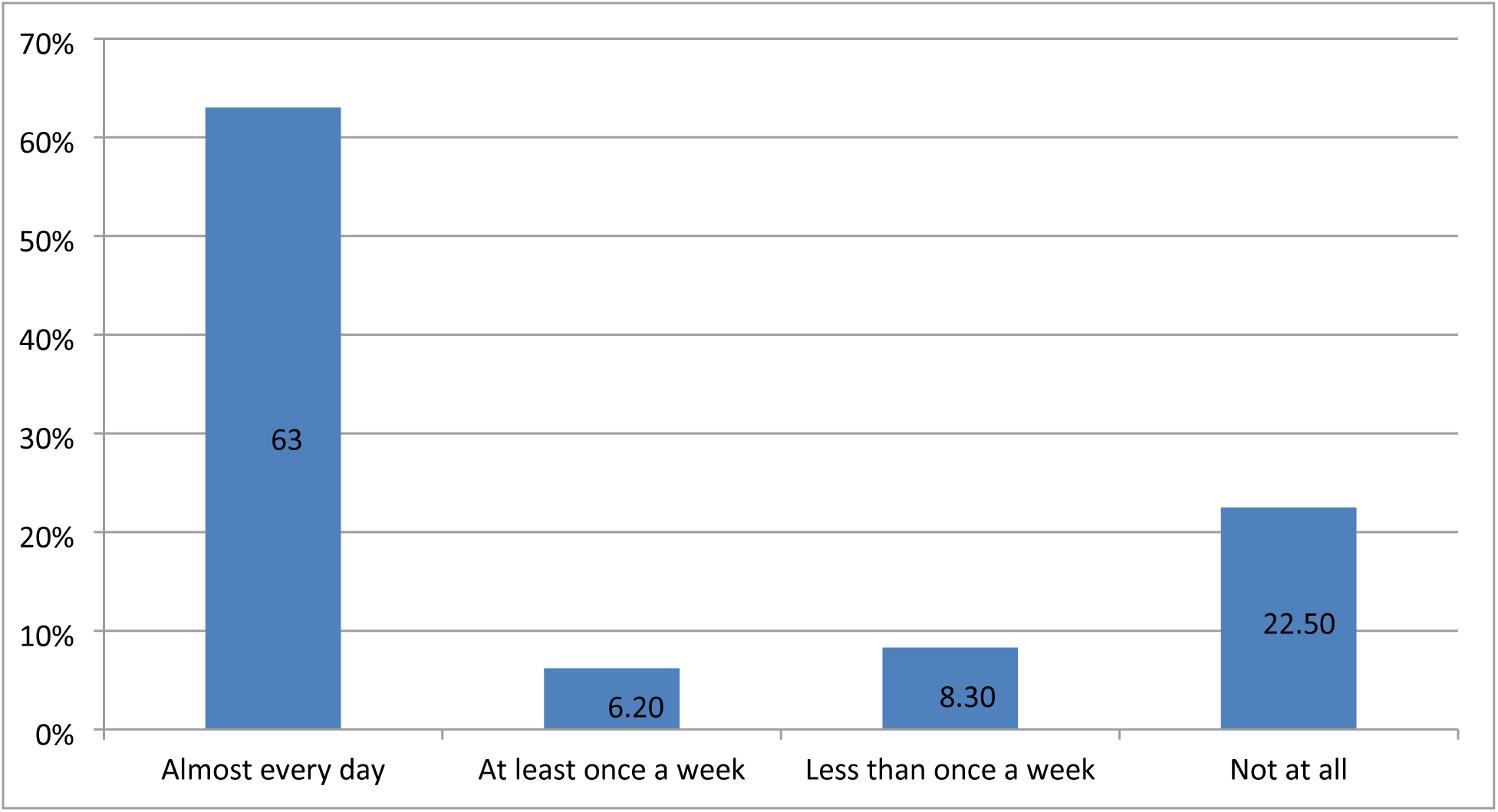
Frequency of watching television among the study participants

### Factors associated with PNC visits

On the bivariate analysis, the independent variables that showed associations with PNC services, including place of delivery, husbands’ income, reading newspapers, ever attending school, watching television, and the adequacy of ANC visits, were statistically significant. These independent variables were again tested together using multivariate analysis to avoid the effects of confounding factors. Before the multivariate analysis was conducted, those variables that were thought to affect PNC services were checked on binary logistic regression using bivariate analysis (see Table 4).

**Table 4:**
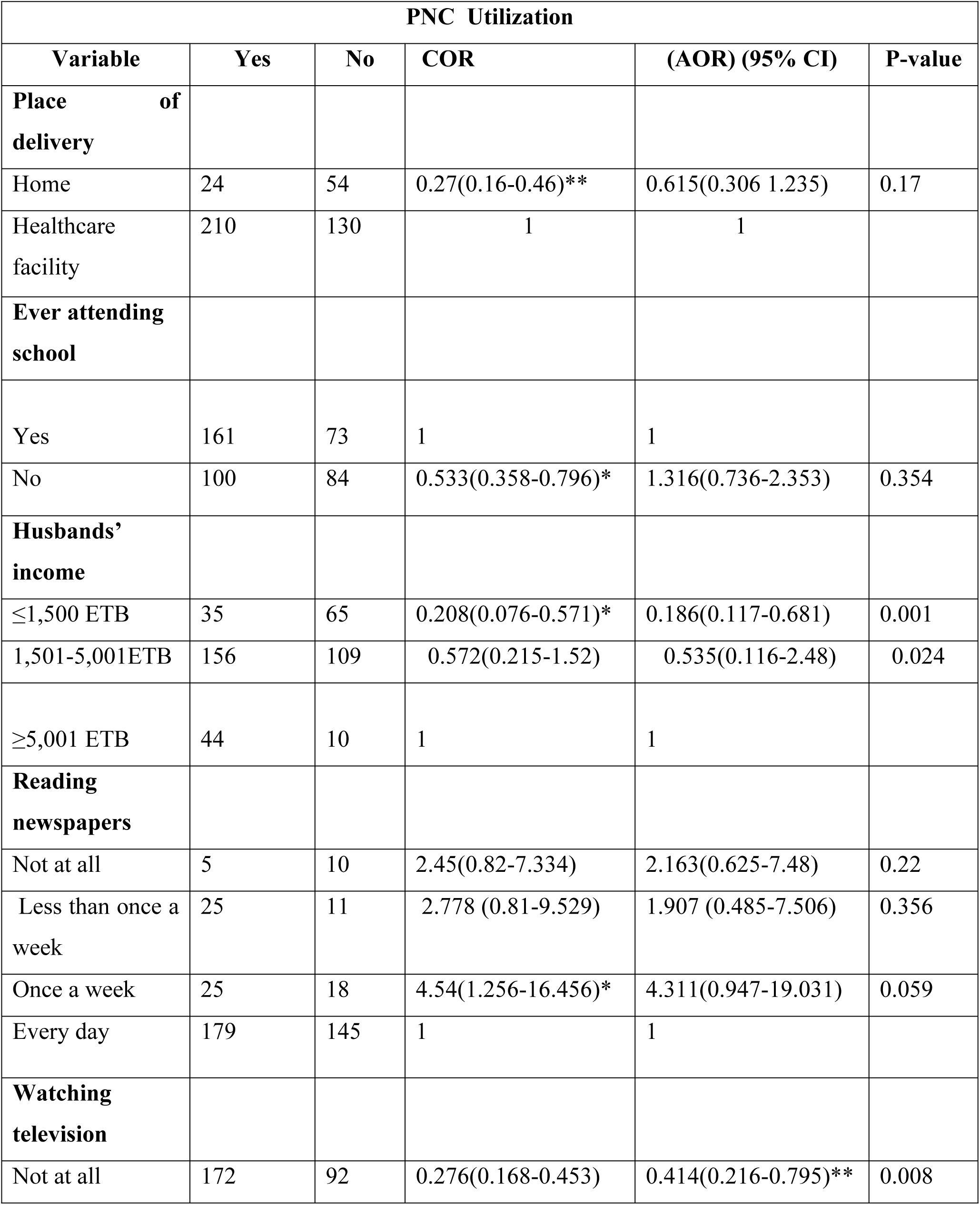
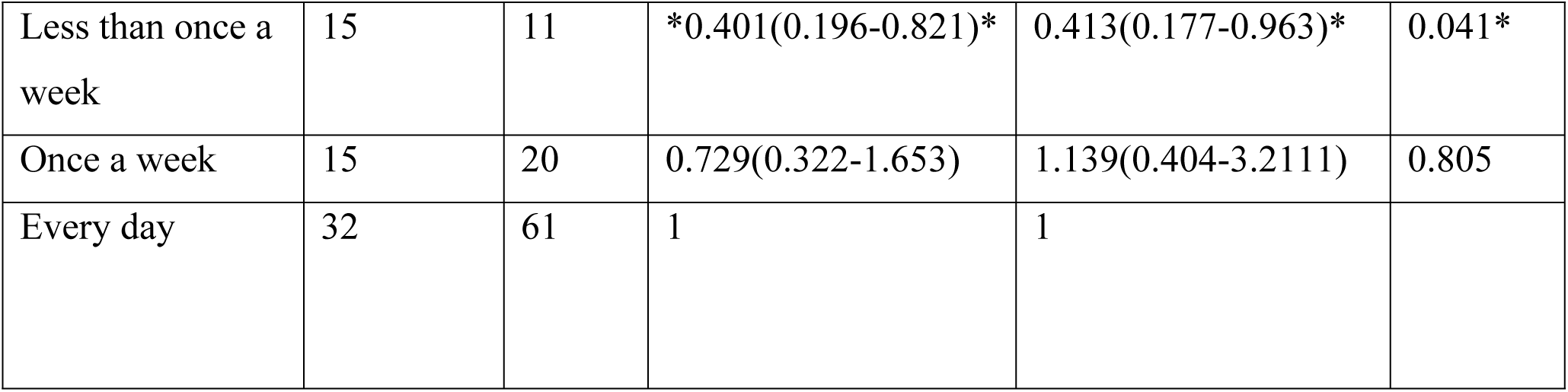
Factors associated with PNC visits of mothers in Awash Fentale District, 2020.

The findings of this research study revealed that women whose husbands had an income of≤1,500 ETB were 81.4% less likely to use PNC services compared to women whose husbands’ income was above 5,001 ETB, with an AOR of 0.186 and p-value of 0.001. A similar statistical association was also seen among women whose husbands’ income was 1,501-5,001 ETB, with an AOR of 0.383 and p-value of 0.024, which is less than the cut-off point of p=0.05. Therefore, regardless of the level or degree of income, husbands’ income was statistically significant and earning less income among husbands might lead women to have less PNC services utilization.

## Discussion

Although there is no sufficient literature available that deals with husbands’ income as a factor that influences PNC services, a study conducted in Swaziland revealed that women in the rich quintile used PNC services more compared to those in the middle wealth quintile (4). A similar research finding in Kenya showed that women from households earning more than US$1 a day were six times more likely to seek PNC services within two days after delivery compared to those women from households earning US$1 and less (5).

Another independent variable that showed statistical significance was watching television. Women who never watched television were 58% less likely to use PNC services compared to women who watched television every day (AOR=0.414 and p-value=0.008), which is less than p=0.05. Women who watched television less than once a week were also 58.7% less likely to use PNC services compared to those women who watched television every day (AOR=0.0413 and p-value=0.041). Therefore, not watching television at all and watching television less than once a week were statistically associated with PNC services utilization. However, a study conducted in Tigray suggested that women who received information about PNC services from health extension workers and a midwife/nurse were 24.87 and 37 times more likely to utilize PNC services respectively compared to women who obtained information from other sources (6). Since obtaining information from healthcare workers was not considered as a factor of PNC services utilization in this study, it would be difficult to compare and contrast with the current findings. Despite the fact that the study conducted in Afghanistan excluded PNC, the findings revealed that media exposure (watching television) was strongly associated with both ANC and SBA utilization (7).

### Women’s Satisfaction with maternal health care services

Client satisfaction is considered one of the desired outcomes of health care and it is directly related with the utilization of health care services, as it reflects the gap between the expected and the actual experience of the service from the client’s point of view (8).

Women who had either PNC visits were asked whether they were satisfied with the care they had received. In this study, 200 (52%) of the respondents were satisfied with the care and attention they received from the health care provider. A similar study conducted in Gamo Gofa Zone showed that 76% of the women were satisfied with the care they received from the health care provider (9). The reason for this higher degree of satisfaction in Gamo Gofa was thought to be the proximity of the health care facility to the women seeking maternal health care. The finding of this study is much lower than the findings in the literature. Long distance and long waiting time were identified as barriers for maternal healthcare services utilization by healthcare workers in Awash Fentale woreda, which may underestimate the overall satisfaction.

Another reason may be because Awash Fentale woreda is located in a pastoralist region, therefore the possibility of accessing skilled labor to give appropriate care is difficult. The healthcare institutions might therefore sometimes be forced to provide the services with under qualified healthcare workers.

Regarding the attitude of health care personnel, 153 (39.7%) of the respondents were satisfied with the attitude of the health care personnel in providing maternal healthcare services. A study conducted in Debre Markos showed that only 33.5% of women were satisfied with the care provided. The most frequently cited reason for dissatisfaction was health care worker-related attitudes, followed by lack of knowledge (10).

Less than half (n=170; 44%) of the participants claimed that they were satisfied with the cleanliness of the health care facility. A study conducted in Nigeria indicated that 82.5% of women were satisfied with the cleanliness of the health care facility (8). In this study, the percentage of satisfaction with regard to cleanliness was very low as compared to the literature. The variation may be because of a real difference in the type of health care facility or differences in the expectations of women.

The participants were also asked about their satisfaction pertaining to the amount of privacy they had and the medication they were provided. Only 135 (35%) of the respondents were satisfied with privacy they had. A study conducted in Debre Markos indicated that approximately 98% of the participants were satisfied with the assurance of privacy (11).The difference might be attributed to the fact that the study in Debre Markos was conducted in a referral teaching hospital where there were a relatively sufficient number of health care professionals and better diagnosis facilities.

In this study, only 82 (21.3%) of the women were satisfied with the medication they were provided. A study conducted in Debre Markos revealed that 73% and 57% of the respondents were satisfied with the explanation of healthcare providers about the drugs prescribed and their side effects respectively (11). A similar study conducted in Assela Hospital in Ethiopia indicated that 87.4% of the mothers were satisfied with the ordered drugs and medical supplies (12).

The findings of this study has a remarkable difference compared to the above mentioned studies. The differences may be attributed to the type of health care facility that was selected during the study. Unlike in Assela and Debre Markos, the majority of the study participants in Awash Fentale woreda received maternal health care services in health care centers. Hence, this difference might influence the level of satisfaction of the participants.

## Conclusion

In this study the utilization of post natal care in Awash Fentale district was 55.45%. Regardless of the level or degree of income, husbands’ income was statistically significant and earning less income among husbands might lead women to have less PNC services utilization. This research has also found that 43% of women were not satisfied with the care and attention given by the healthcare providers and approximately 52% of them were not satisfied with the cleanliness of the healthcare facilities. Hence, much emphasis should be placed on care and attention and the cleanliness of the healthcare facilities in Awash Fentale woreda.

### Limitation of the study

While interpreting the findings of this research, it will be relevant to consider some of the limitations of the study. It is clear that a cross-sectional study design does not allow making causal inferences about the relationship between ANC, DC, and PNC services and risk factors. The study was also limited to Awash Fentale woreda and the findings might not reflect the situation in other woredas of the Afar National Regional State. Limitations on research assistant’s heat exhaustion, language and sampling bias were also considered.

## Data Availability

The data can be shared to relevant bodies

## List of abbreviation

CSA: Central Statistics Agency
DHS: Demographic Health Statistics
EDHS: Ethiopian Demographic Health Statistics
MMR: Maternal Mortality Rate
NGO: Non-Governmental Organization
PNC: Post natal Care
SDG: Sustainable Development Goals
UN: United Nations
WHO: World Health Organization Declaration

## Authors’ Contributions

GW involved in designing of the study, data collection, data analysis, drafting and critically reviewing the manuscript. Likewise, TG involved in critically reviewing the manuscript. All authors read and approved the final manuscript

## Acknowledgements

My special gratitude goes to my supervisor, Prof. Thanyani Gladys Lumadi, for her invaluable guidance and support during the whole process of the research.

The authors would like to thank Yekatit 12 Hospital Medical College for funding this study.

The researchers would also sincerely thank the study participants for their participation in the study.

## Ethics Approval and Consent to Participants

The study protocol was performed in accordance with the ethics principles. Ethical approval was obtained from the institutional review board of University of South Africa. The authors obtained written informed consent from all participants.

## Consent for publication

Consent to publish is not applicable for this manuscript because there is no individual data details like images or videos.

## Availability of Data and Materials

The finding of this study is generated from the data collected and analyzed based on the stated methods and materials. All the data are already found in the manuscript and there are supplementary files. See the additional files.

## REFERENCE

1. Adedoja, O.R., Joshua, A.T. & Olasoji, I. 2020. Awareness and Perception Towards the Utilisation of Primary Healthcare Services in Ado-Ekiti Local Government Area, Ekiti State, Nigeria. Benin Journal of Social Work and Community Development Vol 1, September 2020, pp 23–34

2. Central Statistical Agency (CSA) of Ethiopia. 2016. 2016 Ethiopia Mini-Demographic and Health Survey (EMDHS).Available at: https://dhsprogram.com/pubs/pdf/FR328/FR328.pdf.

3. El Shiekh & Van der Kwaak, A. 2015. Factors Influencing the Utilization of Maternal Healthcare Services by Nomads in Sudan. Pastoralism: Research, Policy and Practice, 2015(5), 23. Available at: 10.1186/s13570-015-0041-x.

4. Atiya, K.M. 2016. Maternal Satisfaction Regarding Quality of Nursing Care During Labor and Delivery in Sulaimani Teaching Hospital. International Journal of Nursing and Midwifery, 8(3), pp. 18–27.

5. Tsawe, M., Moto, A., Netshivhera, T., Relesego, L., Nyathi, C. & Susuman, A.S. 2015. Factors Influencing the Use of Maternal Healthcare Services and Childhood Immunization in Swaziland. International Journal for Equity in Health, 14(32), pp. 1–11.

6. Nzioki, J.M., Onyango, R.O. & Ombaka, J.H. 2017. Socio-Demographic Factors Influencing Maternal and Child Health Service Utilization in Mwingi: A Rural Semi-Arid District in Kenya. American Journal of Public Health Research, 3(1), pp.21–30.

7. Aregay, A., Alemayehu, M., Assefa, H. & Terefe, W. 2014. Factors Associated with Maternal Healthcare Services in Enderta District, Tigray, Northern Ethiopia: A Cross-Sectional Study. American Journal of Nursing Science, 3(6), p.117–125. Available at: http://www.sciencepublishinggroup.com/journal/paperinfo.aspx?journalid=152&doi=10.11648/j.ajns.20140306.15.

8. Shahram, M.S., Hamajima, N. & Reyer, J.A. 2015. Factors Affecting Maternal Healthcare Utilization in Afghanistan: Secondary Analysis of Afghanistan Health Survey 2012. Nagoya Journal of Medical Science, 77(4), pp.595–607.

9. Timane M, Oche D, Umer, Constane K, & Raji,.2017. Awareness and perception towards the utilization of primary health care services in Ado-Ekiti local government area, Ekiti stat, Nigeria. Benin Journal of Social Work and Community Development Vol. 1, September 2020, pp. 23–34

10. Tesfaye, R., Worku, A., Godana, W. & Lindtjorn, B. 2016. Client Satisfaction with Delivery Care Service and Associated Factors in the Public Health Facilities of Gamo Gofa Zone, Southwest Ethiopia: In a Resource-Limited Setting. Obstetrics and Gynecology International, #5798068, 7 pages. Available at: https://www.hindawi.com/journals/ogi/2016/5798068/.

11. Limenih, M.A., Endale, Z.M. & Dachew, B.A., 2016. Postnatal Care Service Utilization and Associated Factors Among Women Who Gave Birth in the Last 12 Months Prior to the Study in Debre Markos Town, Northwestern Ethiopia: A Community-Based Cross-Sectional Study. International Journal of Reproductive Medicine, 2016, #7095352. Available at: https://www.hindawi.com/journals/ijrmed/2016/7095352/.

12. Bitew, K., Ayichiluhm, M. & Yimam, K. 2015. Maternal Satisfaction on Delivery Service and Its Associated Factors Among Mothers Who Gave Birth in Public Health Facilities of Debre Markos Town, Northwest Ethiopia. Biomedical Research International, 2015, pp.1–7.

13. Amdemichael, R., Tafa, M. & Fekadu, H. 2014. Gynecology & Obstetrics Maternal Satisfaction with the Delivery Services in Assela Hospital, Arsi Zone, Oromia Region, Ethiopia. Gynecology Obstetrics, 4(12), 257. Available at: https://www.omicsonline.org/open-access/maternal-satisfaction-with-the-delivery-services-in-assela-hospital-arsi-zone-oromia-region-ethiopia-cas-2161-0932.1000257.php?aid=35831.

